# An Admission-Only Prognostic Model for Early Outcome Prediction After Aneurysmal Subarachnoid Haemorrhage: Development and Temporal Validation

**DOI:** 10.64898/2026.04.28.26352003

**Authors:** Atsushi Nakayashiki, Kunihiko Umezawa, Yasuo Nishijima, Ryutaro Suzuki, Michiko Yokosawa, Hidenori Endo

## Abstract

**Background:** Early prognostic estimation after aneurysmal subarachnoid haemorrhage (aSAH) is often required before complete aneurysm characterisation, neurological reassessment and treatment-related information are available. In this study, we developed and temporally validated an admission-only model for favourable 90-day functional outcomes.

**Methods:** We retrospectively analysed consecutive treatment-eligible patients with aSAH who underwent aneurysm-securing treatment between 2021 and 2025. Patients treated during 2021–2023 and 2024–2025 formed the development and temporal validation cohorts, respectively. Predictors were age, pre-morbid modified Rankin Scale (mRS), admission World Federation of Neurosurgical Societies grade, modified Fisher grade and intracerebral haemorrhage on initial computed tomography. Primary and sensitivity outcomes were 90-day mRS 0–3 and 0–2, respectively. Model performance was assessed using discrimination, Brier score, calibration and decision curve analysis, with contextual benchmarking against admission WFNS grade alone and the SAFIRE grading scale.

**Results:** Overall, 245 patients were included (development cohort, n=160; validation cohort, n=85). In temporal validation, AUCs were 0.868 for mRS 0–3 and 0.870 for mRS 0–2; corresponding Brier scores were 0.134 and 0.147. The admission-only model showed higher discrimination and lower Brier scores than admission WFNS grade alone. The SAFIRE-based contextual benchmark yielded higher performance but required information beyond initial presentation. Exploratory external assessment showed AUCs of 0.790 and 0.775 for 6-month mRS 0–3 and 0–2, respectively.

**Conclusions:** This admission-only model may complement established prognostic scores by supporting early prognostic estimation before all components of the SAFIRE grading scale are available.

**KEY MESSAGES:** *WHAT IS ALREADY KNOWN ON THIS TOPIC:* Aneurysmal subarachnoid haemorrhage is a severe stroke subtype associated with substantial mortality and long-term disability. Existing prognostic scores, including SAFIRE, provide useful risk stratification, but some components may not be uniformly available at the initial presentation before complete aneurysm characterisation and neurological reassessment.

*WHAT THIS STUDY ADDS:* In this study, we developed and temporally validated a simple admission-only prognostic model using five readily available variables: age, premorbid modified Rankin Scale score, admission World Federation of Neurosurgical Societies grade, modified Fisher grade, and intracerebral haemorrhage on initial CT. The model provides individualised probabilities of favourable 90-day functional outcomes before treatment-related information is available, and was evaluated using discrimination, calibration, decision curve analysis, and exploratory external assessment.

*HOW THIS STUDY MIGHT AFFECT RESEARCH, PRACTICE OR POLICY:* By focusing on the earliest clinical time point, this model may help bridge the gap between initial presentation and complete prognostic assessment using established scores.

## INTRODUCTION

Aneurysmal subarachnoid haemorrhage (aSAH) remains one of the most devastating stroke subtypes and is associated with substantial mortality and long-term disability.[1,2] Despite advances in aneurysm treatment and neurocritical care, early prognostic estimation remains challenging,[3] particularly at the initial presentation, when treatment-related information and subsequent clinical events are not yet available. Prognostic tools that can be applied at the earliest clinical time point may support early risk communication, standardised risk stratification, and comparison of case mixes across cohorts.[3,4] Several prognostic models have been developed for aSAH, including the Subarachnoid Hemorrhage International Trialists (SAHIT)[4] model and the SAFIRE[5] grading scale. Although these models provide useful prognostic information and serve as important benchmarks, their complete application may require information that is not uniformly available at the front door of aSAH care. In particular, SAFIRE incorporates aneurysm size, Fisher grade, post-resuscitation World Federation of Neurosurgical Societies (WFNS) grade, and age, and represents a prognostic framework applied after a more complete aneurysm characterisation and neurological reassessment.[5] However, clinicians often need to discuss the prognosis before the treatment modality has been selected and before post-admission factors become available. Moreover, some previously reported models incorporate treatment-related variables or in-hospital complications, limiting their applicability for prognostic assessment at initial presentation.[6,7] The clinical context of care for aSAH has been changing, partly as a result of population ageing, which has become a major issue in Asia, including Japan. Older individuals account for an increasing proportion of patients with stroke, including aSAH.[8–10] Older patients often have more heterogeneous premorbid functional status and recovery potential than younger patients, making admission-based prognostic assessment that accounts for premorbid function particularly important in contemporary treatment-eligible cohorts.[9,11] However, simple temporally validated models that use only the variables available at initial presentation remain limited.

Therefore, in this study, we aimed to develop and temporally validate a simple admission-only prognostic model for favourable 90-day functional outcomes in a contemporary cohort of treatment-eligible patients with aSAH. The model was designed not to replace established scores such as SAFIRE, but to provide a complementary front-door framework for individualised prognostic estimation at initial presentation before complete SAFIRE assessment is available.

## METHODS

### Study design and patient selection

This retrospective single-centre cohort study included consecutive patients with aSAH who underwent aneurysm-securing treatment between January 2021 and December 2025. The source cohort was obtained from the institutional neurosurgical aSAH treatment database. Within this source cohort, no further exclusion criteria were applied to eligible patients. Patients who died before neurosurgical consultation, those without brainstem reflexes who were not considered candidates for aneurysm securing, and those managed entirely outside the neurosurgical treatment pathway were outside the source database and therefore could not be systematically enumerated in this study. Accordingly, the present cohort should be interpreted as a consecutive treatment-eligible and aneurysm-treated aSAH cohort rather than an all-comer aSAH population.

The study was conducted and reported in accordance with the Transparent Reporting of a Multivariable Prediction Model for Individual Prognosis or Diagnosis (TRIPOD) guidelines.[12] The study was approved by the Institutional Review Board, and the requirement for informed consent was waived because of the retrospective study design.

For model development and temporal validation, the study population was divided according to the treatment period; the development and temporal validation cohorts comprised patients treated between 2021 and 2023 and between 2024 and 2025, respectively. Model performance was assessed in the overall cohort and, in an exploratory subgroup analysis, in patients aged ≥ 70 years, who comprised a substantial proportion of the study population.

### Clinical variables and outcome definitions

The admission-only model was defined as a prognostic model using variables available at the initial presentation, before the definitive treatment modality was selected, and before post-admission clinical events occurred. Candidate predictors were selected based on their clinical relevance, availability at the front door of care, and model parsimony. The predictors included age, premorbid modified Rankin Scale (mRS) score, World Federation of Neurosurgical Societies (WFNS) grade, modified Fisher grade, and the presence of intracerebral haemorrhage (ICH) on initial computed tomography (CT).

Age was entered per 10-year increase. Pre-morbid mRS, admission WFNS grade, and modified Fisher grade were dichotomised and entered as binary variables: premorbid mRS ≥ 2, WFNS grade IV–V, and modified Fisher grade 3–4 were coded as 1, and 0 otherwise. ICH was coded as present or absent. The aneurysm size, post-resuscitation WFNS grade, treatment modality, and post-admission events were not included in the admission-only model.

Functional outcome was assessed 90 days after aSAH using the mRS. The primary outcome was favourable functional outcome, defined as a 90-day mRS score of 0–3. Sensitivity outcome was defined as a 90-day mRS score of 0–2. The mRS 0–3 outcome was selected as the primary definition because it represents survival with no more than moderate disability and was considered clinically meaningful in a contemporary treatment-eligible aSAH cohort that included a substantial proportion of older patients.

### Model development

Using the development cohort, we constructed admission-only prognostic models for favourable 90-day functional outcomes. The same pre-specified set of admission variables was used for both outcome definitions. For the primary outcome, defined as an mRS score of 0–3, a multivariable logistic regression model was used. For the sensitivity outcome, defined as mRS 0–2, Firth’s penalised logistic regression was applied because complete separation occurred during model estimation.[13,14] No automated variable-selection procedure was performed. Individual predicted probabilities of favourable outcomes were calculated using the linear predictor derived from the regression coefficients of each model. Detailed equations for calculating the individual predicted probabilities are provided in the online supplemental methods.

### Model performance and temporal validation

The regression coefficients derived from the development cohort were applied without refitting to the temporal validation cohort to estimate the predicted probability of favourable outcome for each patient. Model performance was assessed in the development and temporal validation cohorts, both in the overall population and in patients aged ≥ 70 years. In the subgroup analysis of patients aged ≥ 70 years, the coefficients derived from the overall development cohort were applied without refitting, and the model performance was assessed in an exploratory manner.

Discrimination was assessed using the area under the receiver operating characteristic curve (AUC). The overall prediction accuracy was evaluated using the Brier score. Calibration was assessed using the calibration intercept, calibration slope, and calibration plots, in accordance with established approaches for evaluating prediction model performance.[15,16] Bootstrap bias-corrected 95% confidence intervals (CIs) were calculated for the AUC, Brier score, calibration intercept, and calibration slope using 2000 bootstrap resamples. Decision curve analysis was performed on the temporal validation cohort to assess the potential clinical utility of the admission-only model.[17]

### Model benchmarking and predicted probability stratification

For contextual benchmarking, the admission-only model was compared with admission-WFNS grade alone and a SAFIRE-based benchmark in the temporal validation cohort.[5] The WFNS grade alone was modelled using univariable logistic regression fitted in the development cohort and then applied to the temporal validation cohort without refitting. SAFIRE scores were calculated using a published integer point system. For the Brier score and calibration assessment, the SAFIRE total score was locally calibrated to predicted probabilities in the development cohort and then evaluated in the temporal validation cohort. The admission WFNS and modified Fisher grades were used because the post-resuscitation WFNS and original Fisher grades were not uniformly available; therefore, this analysis should be interpreted as a contextual benchmark rather than a strict replication of the original SAFIRE model.

To assess the clinically interpretable risk stratification, patients in the temporal validation cohort were divided into tertiles according to the admission-only predicted probability of favourable outcomes for each outcome definition. When identical predicted probabilities occurred at a tertile boundary, the tied observations were maintained in the same stratum and not split. These strata were used for descriptive risk stratification and were not prespecified clinical decision thresholds.[18]

### Exploratory external assessment

As an exploratory external assessment, the final admission-only model was applied to an independent published cohort.[19] Details of cohort selection, assumptions and performance assessment are provided in the online supplemental methods.

### Statistical analysis

Continuous variables are presented as mean ± SD, and categorical variables as number (%). Between-group comparisons were performed using Welch’s t-test for continuous variables, Fisher’s exact test for binary categorical variables, and the χ^2^ test for categorical variables with more than two categories. Candidate predictors were prespecified based on clinical relevance, availability at initial presentation, and model parsimony, rather than being selected using automated data-driven procedures. Odds ratios (ORs) with 95% CIs and p-values are reported for the primary and sensitivity models as descriptive measures of association. All statistical tests were two-sided, and p-values < 0.05 were considered statistically significant. All 245 patients had complete data on prespecified admission predictors and 90-day mRS outcomes; therefore, no imputation was performed. Statistical analyses were performed using JMP Student’s Edition 19 (SAS Institute, Cary, NC, USA) and GraphPad Prism 11 (GraphPad Software, San Diego, CA, USA).

## RESULTS

### Baseline characteristics

A total of 245 consecutive patients with treatment-eligible aSAH, who underwent aneurysm-securing treatment between January 2021 and December 2025, were included. Of these, 160 patients treated during 2021–2023 were assigned to the development cohort and 85 patients treated during 2024–2025 were assigned to the temporal validation cohort; 107 patients were aged ≥ 70 years (Figure 1). The mean age was 66.7 ± 15.6 years in the development cohort and 65.7 ± 13.9 years in the validation cohort (Table 1).

**Table 1.** Baseline characteristics of the development and temporal validation cohorts.

| Characteristics | Development<br>(2021–2023)<br>n = 160 | Validation (2024–<br>2025)<br>n = 85 | P-value |
| --- | --- | --- | --- |
| Age, mean ± SD, years | 66.7 ± 15.6 | 65.7 ± 13.9 | 0.633 |
| Age $\geq 70$ years, n (%) | 75 (46.9) | 32 (37.6) | 0.178 |
| Female sex, n (%) | 113 (70.6) | 63 (74.1) | 0.655 |
| Current smoking, n (%) | 55 (34.4) | 31 (36.5) | 0.779 |
| Diabetes mellitus, n (%) | 12 (7.5) | 9 (10.6) | 0.474 |
| Dyslipidaemia, n (%) | 27 (16.9) | 14 (16.5) | 1.000 |
| Hypertension, n (%) | 78 (48.8) | 34 (40.0) | 0.226 |
| Antiplatelet use, n (%) | 10 (6.3) | 3 (3.5) | 0.551 |
| Anticoagulant use, n (%) | 3 (1.9) | 3 (3.5) | 0.421 |
| Pre-morbid mRS distribution, n (%) |  |  | 0.985 |
| Pre-morbid mRS $\geq 2$, n (%) | 15 (9.4) | 9 (10.6) | 0.822 |
| 0 | 141 (88.1) | 73 (85.9) |  |
| 1 | 4 (2.5) | 3 (3.5) |  |
| 2 | 9 (5.6) | 5 (5.9) |  |
| 3 | 3 (1.9) | 2 (2.4) |  |
| 4 | 3 (1.9) | 2 (2.4) |  |
| WFNS grade distribution, n (%) |  |  | 0.083 |
| WFNS grade IV–V, n (%) | 65 (40.6) | 29 (34.1) | 0.337 |
| I | 53 (33.1) | 22 (25.9) |  |
| II | 34 (21.2) | 24 (28.2) |  |
| III | 8 (5.0) | 10 (11.8) |  |
| IV | 42 (26.2) | 14 (16.5) |  |
| V | 23 (14.4) | 15 (17.6) |  |
| Modified Fisher grade distribution, n (%) |  |  | 0.923 |
| Modified Fisher grade 3–4, n (%) | 91 (56.9) | 47 (55.3) | 0.892 |
| 1 | 33 (20.6) | 20 (23.5) |  |
| 2 | 36 (22.5) | 18 (21.2) |  |
| 3 | 22 (13.8) | 13 (15.3) |  |
| 4 | 69 (43.1) | 34 (40.0) |  |
| Intracerebral haemorrhage, n (%) | 48 (30.0) | 20 (23.5) | 0.298 |
| Aneurysm location, n (%) |  |  | 0.171 |
| ACA | 58 (36.3) | 32 (37.6) |  |
| MCA | 34 (21.3) | 26 (30.6) |  |
| ICA | 53 (33.1) | 24 (28.2) |  |
| Posterior circulation | 15 (9.4) | 3 (3.5) |  |
| Dome size, mean ± SD, mm | 6.0 ± 3.2 | 5.7 ± 2.8 | 0.506 |
| Neck size, mean ± SD, mm | 3.3 ± 1.8 | 3.1 ± 1.4 | 0.316 |
| Treatment modality, n (%) |  |  | < 0.001 |
| Open surgery, n (%) | 39 (24.4) | 45 (52.9) |  |
| Endovascular treatment, n (%) | 121 (75.6) | 40 (47.1) |  |
| Favourable outcome at 90 days, n (%) |  |  |  |
| mRS 0–3, n (%) | 98 (61.3) | 61 (71.8) | 0.122 |
| mRS 0–2, n (%) | 85 (53.1) | 54 (63.5) | 0.137 |

**Notes:** Values are presented as mean ± SD or n (%). P-values were calculated using Welch’s t-test for continuous variables, Fisher’s exact test for binary categorical variables, and the $\chi^2$ test for categorical variables with more than two categories. ACA: Anterior cerebral artery, ICA: Internal carotid artery, ICH: Intracerebral haemorrhage, MCA: Middle cerebral artery, mRS: Modified Rankin Scale, SD: Standard deviation, WFNS: World Federation of Neurosurgical Societies.

**Figure 1.**
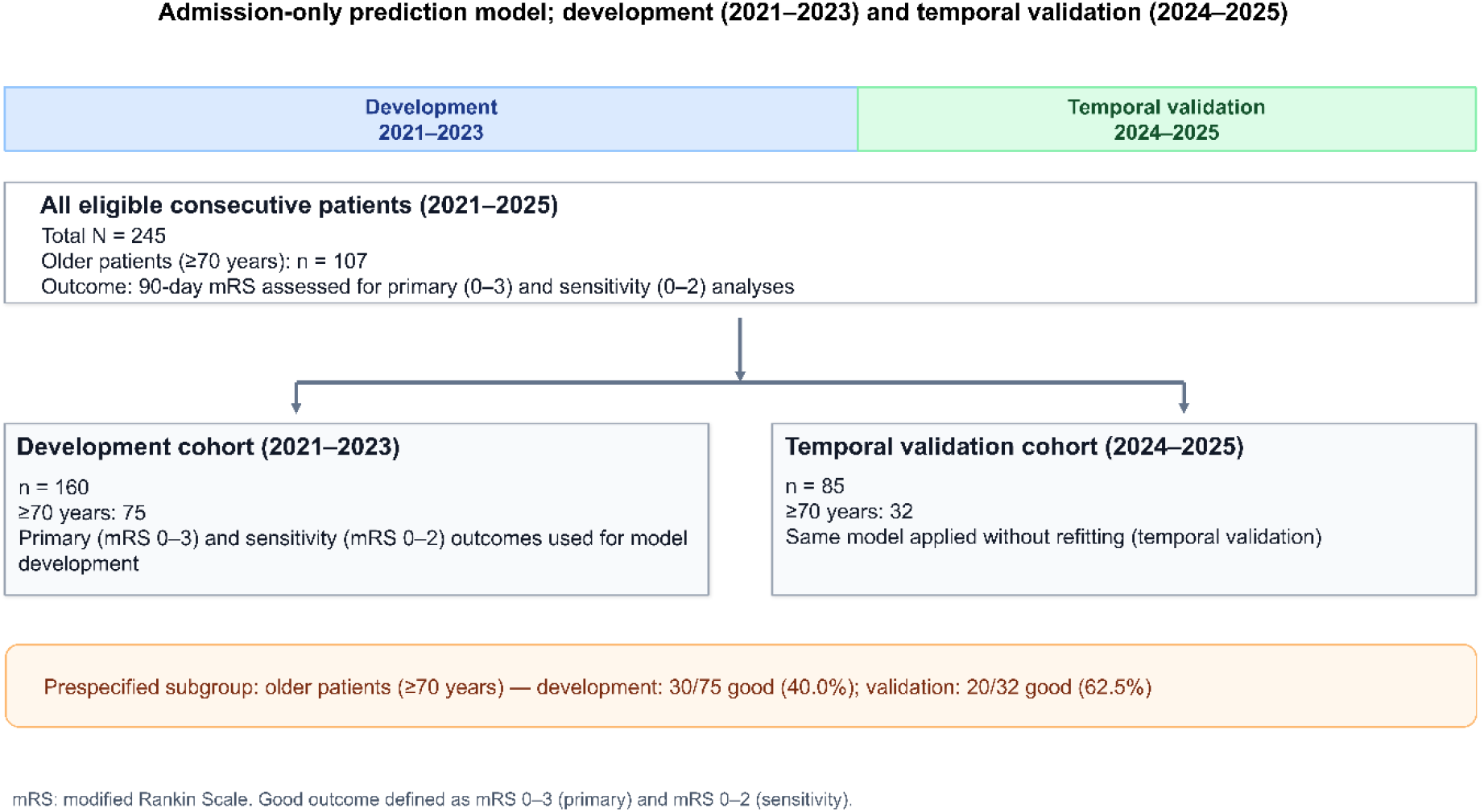
Study flow and cohort allocation for model development and temporal validation. In total, 245 consecutive treatment-eligible patients with aneurysmal subarachnoid haemorrhage who underwent aneurysm-securing treatment were included in the study. Patients treated during 2021–2023 comprised the development cohort, and those patients treated during 2024–2025 comprised the temporal validation cohort. No further exclusion criteria were applied to the source treatment database. aSAH: Aneurysmal subarachnoid haemorrhage, mRS: Modified Rankin Scale.

The distributions of the premorbid mRS score, WFNS grade, modified Fisher grade, and 90-day functional outcomes are shown in Table 1. Overall, the baseline characteristics were generally comparable between the cohorts. The treatment modality differed between the cohorts, with endovascular treatment performed more frequently in the development cohort and open surgery performed more frequently in the validation cohort. Favourable outcomes at 90 days were observed in 98 of 160 patients (61.3%) in the development cohort and 61 of 85 patients (71.8%) in the validation cohort for mRS 0–3, and in 85 of 160 patients (53.1%) and 54 of 85 patients (63.5%), respectively, for mRS 0–2. Detailed baseline characteristics of patients aged ≥ 70 years are presented in Table S1.

### Model development and discrimination

Final admission-only models for predicting favourable 90-day functional outcomes were developed in the development cohort, and the regression coefficients are presented in Table 2. In the primary model for mRS 0–3, older age, premorbid mRS ≥ 2, WFNS grade IV–V, and modified Fisher grade 3–4 were associated with a lower likelihood of favourable outcome, whereas ICH showed no clear association. In the sensitivity model for mRS 0–2, older age, premorbid mRS ≥ 2, and WFNS grade IV–V showed similar associations. The full ORs, 95% CIs, and p-values are provided in Table S2.

**Table 2.** Regression coefficients and variable coding of the admission-only prognostic model.

| Predictor | Coding/unit | mRS 0–3 $\beta$ | mRS 0–2 Firth $\beta$ |
| --- | --- | --- | --- |
| Intercept | — | 5.829425 | 5.983238 |
| Age | Per 10-year increase | –0.436149 | –0.596888 |
| Pre-morbid mRS | $\geq 2$ vs. 0–1 | –2.059763 | –2.924046 |
| WFNS grade | IV–V vs. I–III | –3.096286 | –2.986462 |
| Modified Fisher grade | 3–4 vs. 1–2 | –1.098374 | –0.808869 |
| ICH | Present vs. absent | –0.123058 | –0.066414 |

**Notes:** Predicted probabilities were calculated as
where LP denotes the linear predictor. Age was entered per 10-year increase. Binary variables were coded as 1 when present and 0 otherwise. ICH: Intracerebral haemorrhage, mRS: Modified Rankin Scale, WFNS: World Federation of Neurosurgical Societies.

In the temporal validation cohort, the admission-only model maintained good discrimination, with AUCs of 0.868 and 0.870, and corresponding Brier scores of 0.134 and 0.147 for mRS 0–3 and mRS 0– 2, respectively. Compared with the WFNS grade alone, the admission-only model showed higher discrimination and lower Brier scores for both outcome definitions. The SAFIRE-based benchmark showed higher AUCs and lower Brier scores than the admission-only model but was considered a contextual benchmark because complete SAFIRE assessment requires information not uniformly available at initial presentation. The temporal validation and benchmark results are presented in Table 3.

**Table 3.** Temporal validation and benchmarking against established scores.

| Model | AUC<br>(mRS 0–3/0–2) | Brier<br>(mRS 0–3/0–2) | Clinical timing |
| --- | --- | --- | --- |
| Admission-only model | 0.868/0.870 | 0.134/0.147 | Initial presentation |
| WFNS grade alone | 0.834/0.820 | 0.155/0.179 | Initial presentation |
| SAFIRE-based benchmark | 0.895/0.905 | 0.125/0.127 | After aneurysm<br>characterisation |

**Notes:** Information required by the model: Admission-only model—age, premorbid mRS, admission WFNS, modified Fisher grade, ICH; WFNS grade alone—admission WFNS grade; SAFIRE-based benchmark — age, aneurysm size, Fisher/mFisher grade, rWFNS/admission WFNS (assessed after aneurysm characterisation and neurological reassessment). The SAFIRE-based benchmark used the published integer point system for AUC. For Brier score and calibration assessment, the SAFIRE total score was locally calibrated to predicted probabilities in the development cohort and evaluated in the temporal validation cohort. Because post-resuscitation WFNS and original Fisher grades were not uniformly available, admission WFNS and modified Fisher grades were used; therefore, this analysis should be interpreted as a contextual benchmark rather than a strict replication of the original SAFIRE model. AUC: Area under the receiver operating characteristic curve, ICH: Intracerebral haemorrhage, mRS: Modified Rankin Scale, WFNS: World Federation of Neurosurgical Societies.

In the exploratory subgroup analyses of patients aged ≥ 70 years, the admission-only model retained acceptable discrimination. In the temporal validation cohort, the AUCs were 0.842 and 0.844 for mRS scores of 0–3 and 0–2, respectively. Detailed performance metrics, including bootstrap bias-corrected 95% CIs, Brier scores, and calibration measures, are provided in Table S3 and the receiver operating characteristic curves are shown in Figure S1.

### Calibration performance

Calibration plots for the temporal validation cohort are shown in Figure 2, and detailed calibration metrics are provided in Table S3. In the overall validation cohort, the primary model for mRS 0–3 showed a Brier score of 0.134, calibration intercept of 0.587, and calibration slope of 0.827. For the sensitivity model for an mRS score of 0–2, the corresponding values were 0.147, 0.546, and 0.787, respectively.

**Figure 2.**
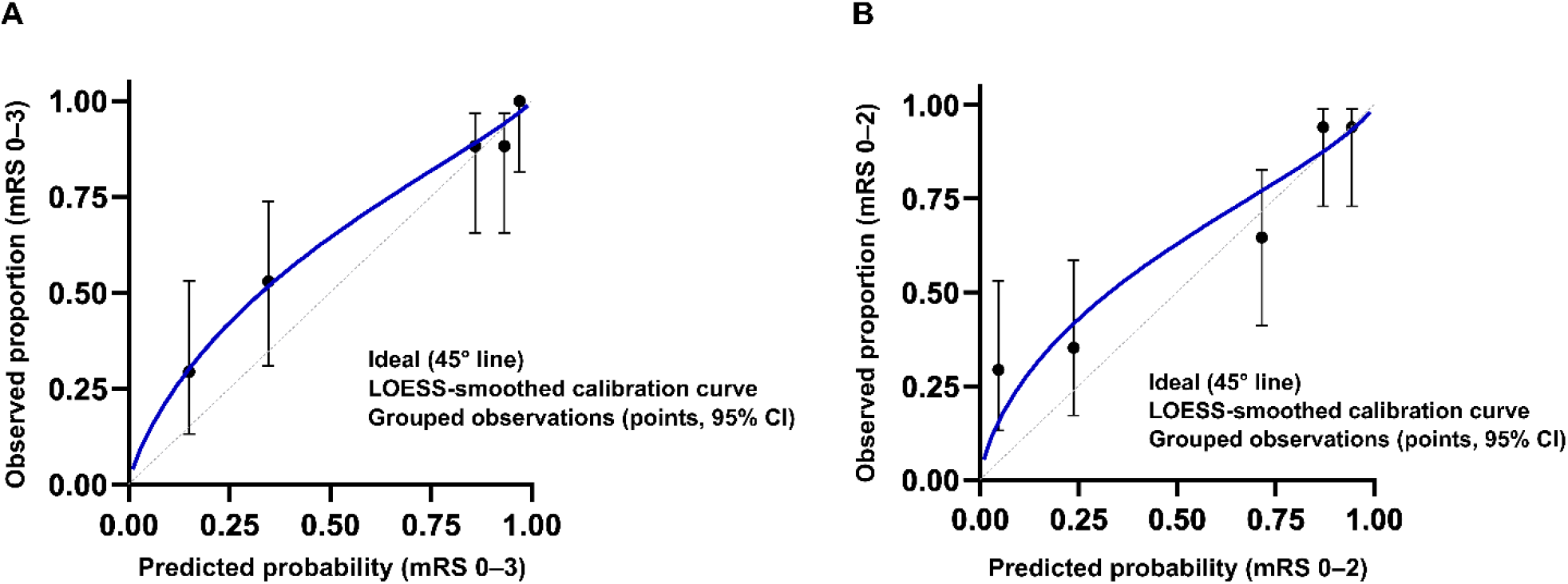
Calibration plots of the admission-only prognostic model in the temporal validation cohort. Calibration plots are shown for favourable 90-day functional outcomes, defined as mRS 0–3 (A) and mRS 0–2 (B). Points indicate observed outcome rates grouped according to quintiles of predicted probability, and error bars indicate Wilson’s 95% confidence intervals. The solid curve represents the fitted calibration curve, and the dashed diagonal line indicates perfect calibration. For mRS 0–3, the Brier score, calibration intercept, and calibration slope were 0.134, 0.587, and 0.827, respectively. For mRS 0–2, the corresponding values were 0.147, 0.546, and 0.787. mRS: Modified Rankin Scale.

In patients aged ≥ 70 years, the calibration estimates were less stable, reflecting the smaller subgroup size. In the temporal validation cohort, the Brier score, calibration intercept, and calibration slope were 0.169, 0.446 and 0.673 for mRS 0–3, and 0.192, 0.381 and 0.577 for mRS 0–2, respectively. Overall, the calibration was generally acceptable in the validation cohort, whereas predictions in older patients, particularly for stricter mRS 0–2 outcomes, should be interpreted with caution. The positive calibration intercepts in the validation cohort and the higher observed-than-predicted event rate in the low-predicted-probability stratum suggest that the model tended to underestimate favourable outcomes in some patients. This may partly reflect differences in baseline outcome rates and institutional practice between the development and validation periods and indicates that recalibration may be required before clinical implementation.

### Predicted probability strata

Patients in the temporal validation cohort were stratified into tertiles according to the admission-only predicted probability of a favourable outcome for each outcome definition (Table 4). For mRS 0–3, the observed favourable outcome rates increased across the low-, intermediate-, and high-predicted-probability strata: 35.7% (10/28), 82.8% (24/29), and 96.4% (27/28), with corresponding mean predicted probabilities of 19.9%, 79.3%, and 95.7%, respectively.

**Table 4.** Observed outcomes across admission-only predicted probability strata in the temporal validation cohort.

| Stratum | n (mRS 0–3/0–2) | mRS 0–3 | mRS 0–2 |
| --- | --- | --- | --- |
|  |  | Predicted %/Observed | Predicted %/Observed |
| Low | 28/28 | 19.9/35.7% (10/28) | 11.6/21.4% (6/28) |
| Intermediate | 29/28* | 79.3/82.8% (24/29) | 65.7/75.0% (21/28) |
| High | 28/29* | 95.7/96.4% (27/28) | 91.5/93.1% (27/29) |

**Notes:** Predicted probability strata were defined by the tertile cutoff points of the admission-only predicted probability within the temporal validation cohort. When identical predicted probabilities occurred at the tertile boundary, the tied observations were maintained within the same stratum. Strata were used for descriptive risk stratification and were not prespecified clinical decision thresholds. \*The number of patients in the strata differed between outcomes because strata were separately defined for each outcome-specific predicted probability and because tied predicted probabilities were not split across strata.

For mRS 0–2, the observed favourable outcome rates increased similarly across the strata: 21.4% (6/28), 75.0% (21/28), and 93.1% (27/29), with corresponding mean predicted probabilities of 11.6%, 65.7%, and 91.5%, respectively. These findings support the descriptive ability of the admission-only model to stratify patients into clinically interpretable prognostic groups, although the strata were not pre-specified clinical decision thresholds.

### Decision curve analysis

Decision curve analysis was performed in the temporal validation cohort (Figure 3). For both the mRS 0–3 and mRS 0–2 outcomes, the admission-only model showed greater net benefit than the classify-none strategy across the evaluated threshold probabilities. Compared with the classify-all strategy and WFNS grade alone, the admission-only model showed potential clinical utility across parts of the clinically relevant threshold range, particularly in the intermediate-to-higher threshold range. These findings suggest that the admission-only model may provide additional value for early risk stratification beyond the admission WFNS grade alone, although the results should be interpreted as supportive rather than definitive evidence of clinical utility.

**Figure 3.**
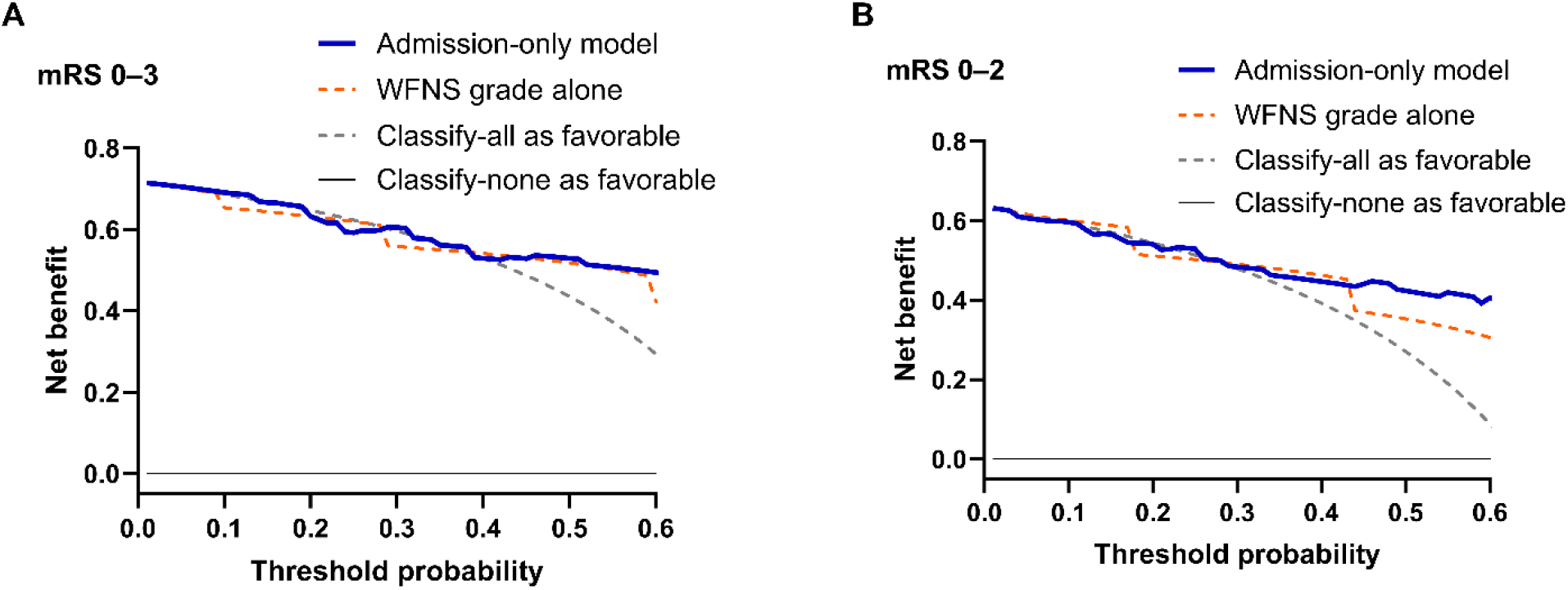
Decision curve analysis of the admission-only prognostic model in the temporal validation cohort. Decision curve analysis was performed for favourable 90-day functional outcomes, defined as mRS 0–3 (A) and mRS 0–2 (B). The net benefit was plotted across a range of threshold probabilities. The admission-only model was compared with admission WFNS grade alone and the default strategies of classifying all or no patients as having a favourable outcome. DCA: Decision curve analysis, mRS: Modified Rankin Scale, WFNS: World Federation of Neurosurgical Societies.

### Exploratory external assessment

Results of the exploratory external assessment are summarised in the online supplemental results, Table S4 and Figure S2. In brief, the AUCs were 0.790 for 6-month mRS 0–3 and 0.775 for 6-month mRS 0– 2.

## DISCUSSION

In this study, we developed and temporally validated a simple admission-only prognostic model for favourable 90-day functional outcomes in treatment-eligible patients with aSAH. The model showed good discrimination in the temporal validation cohort for both mRS 0–3 and mRS 0–2 outcomes and performed better than admission WFNS grade alone, whereas the SAFIRE-based benchmark showed numerically higher performance as a contextual comparator. Calibration was generally acceptable in the overall validation cohort but less stable in older patients, particularly for mRS 0–2.[15] Decision curve analysis and predicted-probability strata supported the potential usefulness of the model for early risk stratification.[17,18] Exploratory external assessment showed supportive discrimination despite important assumptions regarding pre-morbid mRS and outcome timing.[19]

A key feature of the proposed model is the clinical timepoint at which it is intended for use. Several prognostic models for aSAH, including SAHIT and SAFIRE, have been reported and remain important benchmarks.[4,5] SAFIRE is a practical and well-established scoring system, but its complete application requires information such as aneurysm size, Fisher grade, and post-resuscitation WFNS grade. However, in routine practice, clinicians are often required to provide an initial estimate of the prognosis before complete aneurysm characterisation, neurological reassessment, and treatment-related information are available. The model was designed to address this earlier clinical window; therefore, it should not be interpreted as a replacement for SAFIRE but rather as a complementary front-door framework for individualised prognostic estimation before complete SAFIRE assessment is available. The SAFIRE-based benchmark was applied as a contextual comparator using the admission WFNS and modified Fisher grades because the post-resuscitation WFNS and original Fisher grades were not uniformly available. Therefore, this comparison should be interpreted as an assessment of the variable composition in an early clinical time window rather than a strict replication of the original SAFIRE model. The higher performance of the SAFIRE-based benchmark, likely reflecting at least in part the inclusion of aneurysm size, reinforces that the admission-only model should be viewed as complementary rather than a replacement.

This model has practical advantages for early risk communication because it uses five readily available variables: age, pre-morbid mRS, admission WFNS grade, modified Fisher grade and ICH on initial CT. These variables are usually available before definitive treatment selection and post-admission events. The model therefore enables calculation of individual predicted probabilities at admission, which may support family communication, standardised risk stratification and comparison of baseline case mix.[18] However, because the cohort consisted of treatment-eligible patients who underwent aneurysm-securing treatment, the model should not be used to determine treatment eligibility, treatment withholding or withdrawal decisions.[20]

These findings should also be interpreted in the context of the contemporary aging population with aSAH. Older patients accounted for a substantial proportion of the cohort, and premorbid functional status was incorporated as an admission variable. This is clinically relevant because older patients often have more heterogeneous baseline functions and recovery potentials than younger patients. In the exploratory subgroup analysis of patients aged ≥ 70 years, the model retained acceptable discrimination, but the calibration estimates were less stable, particularly for mRS scores of 0–2.[15,16] This likely reflects both the heterogeneity of the older patients and the limited sample size of the older validation subgroup.[21,22] Therefore, the present model should not be regarded as an elderly-specific prognostic model and predictions in older patients should be interpreted with caution.

Moreover, importantly, the model was evaluated across a temporal shift in institutional treatment practice. During the study period, the study institution changed from an endovascular treatment-first default strategy to a more modality-neutral, individualised upfront treatment selection pathway. Endovascular treatment was more frequently performed in the development cohort, whereas open surgery was more frequently performed in the temporal validation cohort. Despite this shift, the admission-only model retained good discrimination in the validation cohort. This finding supports the potential robustness of a model that does not require treatment-related information at the time of prediction. However, this should not be interpreted as evidence that treatment modality has no prognostic effect. Rather, the model is intended to provide prognostic estimates before the treatment modality and post-admission clinical course are known.

The exploratory external assessment provided supportive but limited evidence for generalisability. Because pre-morbid mRS was unavailable and 6-month rather than 90-day mRS was used, this analysis should not be regarded as formal external validation. Further multicentre validation using harmonised 90-day outcomes and complete admission variables is required before clinical implementation.[23]

This study has some limitations. First, this was a retrospective single-centre study, and the cohort was restricted to treatment-eligible patients who underwent aneurysm-securing treatment. Patients who died before neurosurgical consultation or were managed outside the neurosurgical treatment pathway could not be systematically enumerated, and the results cannot be generalised to an all-comer aSAH population. Second, although temporal validation was performed, formal multicentre external validation is necessary. Third, the older subgroup, particularly in the temporal validation cohort, was small, resulting in uncertainty in the calibration estimates. Fourth, the SAFIRE-based analysis was a contextual benchmark rather than a strict replication of the original SAFIRE model because the post-resuscitation WFNS grade and original Fisher grade were not uniformly available. Fifth, ICH was retained as a pre-specified predictor because it reflects haemorrhage burden on initial CT; its modest independent contribution may reflect overlap with the WFNS and modified Fisher grades. Finally, by design, the model excluded aneurysm size, treatment modality, and post-admission events. This may limit the maximal predictive accuracy but preserves the intended role of the model as a simple admission-only framework for prognostic estimation at initial presentation.

## CONCLUSION

The admission-only model developed in this study provided individualised estimates of favourable 90-day functional outcomes in a contemporary cohort of treatment-eligible patients with aSAH. By using variables available at the initial presentation, the model may support early risk communication and standardised risk stratification before complete SAFIRE assessment is available. The model is not intended to replace established prognostic scores and further multicentre external validation is required before clinical implementation.

## Data availability disclosure

The data supporting the findings of this study are available from the corresponding author upon request.

