## Supplementary material for "An Admission-Only Prognostic Model for Early Outcome Prediction After Aneurysmal Subarachnoid Haemorrhage: Development and Temporal Validation": Supplymentary Material

**Online Supplemental Material**

### Supplemental Methods

#### Calculation of individual predicted probabilities

Predicted probabilities of favourable outcome were calculated using：

$$\mathrm{P} = \frac{1}{1 + e^{-LP}}$$

where P denotes the predicted probability and LP denotes the linear predictor.

For the primary model (90-day mRS 0–3), LP = 5.829425 − 0.436149 × (age per 10-year increase) − 2.059763 × (pre-morbid mRS ≥ 2) − 3.096286 × (WFNS grade IV–V) − 1.098374 × (modified Fisher grade 3–4) − 0.123058 × (ICH present).

For the sensitivity model (90-day mRS 0–2), LP = 5.983238 − 0.596888 × (age per 10-year increase) − 2.924046 × (pre-morbid mRS ≥ 2) − 2.986462 × (WFNS grade IV–V) − 0.808869 × (modified Fisher grade 3–4) − 0.066414 × (ICH present).

Age was entered per 10-year increase. Binary variables were coded as 1 for pre-morbid mRS ≥ 2, WFNS grade IV–V, modified Fisher grade 3–4, and ICH present, and as 0 otherwise. The mRS 0–2 model was fitted using Firth's penalised logistic regression.

#### Model performance assessment

Discrimination was evaluated using the area under the receiver operating characteristic curve (AUC). The overall prediction accuracy was summarised using the Brier score. Calibration was assessed using calibration intercepts and slopes estimated by logistic recalibration of the observed outcome on the logit of the predicted probability. Bootstrap bias-corrected 95% confidence intervals (CIs) were calculated for the model performance metrics using 2000 bootstrap resamples.

For subgroup analyses in patients aged ≥ 70 years, coefficients derived from the overall development cohort were applied without refitting, and performance was assessed in the development and temporal validation cohorts.

**Exploratory external assessment**

For the exploratory external assessment, the final admission-only model coefficients were applied to an independent published cohort. Patients without available 6-month mRS data and those with missing modified Fisher grade were excluded. Because pre-morbid mRS was unavailable, all patients were assumed to have pre-morbid mRS < 2. The 6-month mRS was used because the 90-day mRS was unavailable; therefore, this analysis was considered exploratory external assessment rather than formal external validation. Model performance was assessed using the AUC, Brier score, calibration intercept and calibration slope.

**Supplemental Results**

After excluding 31 patients without available 6-month mRS data and one patient with a missing modified Fisher grade, 198 patients were included in the exploratory external assessment. The model yielded AUCs of 0.790 for 6-month mRS 0–3 and 0.775 for 6-month mRS 0–2. The corresponding Brier scores were 0.194 and 0.202, respectively. Calibration intercepts and slopes are presented in Table S4.

### Online supplemental Tables

### Table S1. Detailed baseline characteristics in patients aged ≥ 70 years

| Characteristics | Development cohort (2021–2023, n = 75) | Validation cohort (2024–2025, n = 32) | P-value |
|---|---|---|---|
| Age, mean ± SD, years | 80.7 ± 7.4 | 80.3 ± 6.6 | 0.832 |
| Female sex, n (%) | 55 (73.3) | 28 (87.5) | 0.133 |
| Current smoking, n (%) | 17 (22.7) | 5 (15.6) | 0.602 |
| Diabetes mellitus, n (%) | 11 (14.7) | 4 (12.5) | 1.000 |
| Dyslipidaemia, n (%) | 19 (25.3) | 6 (18.8) | 0.619 |
| Hypertension, n (%) | 48 (64.0) | 14 (43.8) | 0.058 |
| Antiplatelet use, n (%) | 9 (12.0) | 2 (6.2) | 0.500 |
| Anticoagulant use, n (%) | 3 (4.0) | 2 (6.2) | 0.634 |
| Pre-morbid mRS distribution, n (%) | | | 0.876 |
| 0 | 57 (76.0) | 22 (68.8) | |
| 1 | 4 (5.3) | 2 (6.2) | |
| 2 | 9 (12.0) | 4 (12.5) | |
| 3 | 3 (4.0) | 2 (6.2) | |
| 4 | 2 (2.7) | 2 (6.2) | |
| Pre-morbid mRS ≥ 2, n (%) | 14 (18.7) | 8 (25.0) | 0.448 |
| WFNS grade distribution, n (%) | | | 0.122 |
| I | 16 (21.3) | 8 (25.0) | |
| II | 13 (17.3) | 9 (28.1) | |
| III | 4 (5.3) | 5 (15.6) | |
| IV | 25 (33.3) | 5 (15.6) | |
| V | 17 (22.7) | 5 (15.6) | |
| WFNS grade IV–V, n (%) | 42 (56.0) | 10 (31.2) | 0.022 |

| | | | |
|---|---|---|---|
| Modified Fisher grade distribution, n (%) | | | 0.211 |
| 1 | 9 (12.0) | 7 (21.9) | |
| 2 | 18 (24.0) | 10 (31.2) | |
| 3 | 6 (8.0) | 4 (12.5) | |
| 4 | 42 (56.0) | 11 (34.4) | |
| Modified Fisher grade 3–4, n (%) | 48 (64.0) | 15 (46.9) | 0.133 |
| Intracerebral haemorrhage, n (%) | 30 (40.0) | 7 (21.9) | 0.080 |
| Aneurysm location, n (%) | | | 0.695 |
| ACA | 24 (32.0) | 11 (34.4) | |
| MCA | 19 (25.3) | 11 (34.4) | |
| ICA | 26 (34.7) | 8 (25.0) | |
| Posterior circulation | 6 (8.0) | 2 (6.2) | |
| Dome size, mean ± SD, mm | 6.9 ± 3.7 | 5.9 ± 3.1 | 0.171 |
| Neck size, mean ± SD, mm | 3.9 ± 2.2 | 3.2 ± 1.6 | 0.069 |
| Treatment modality, n (%) | | | 0.182 |
| Open surgery, n (%) | 22 (29.3) | 14 (43.8) | |
| Endovascular treatment, n (%) | 53 (70.7) | 18 (56.2) | |
| Favourable outcome at 90 days, n (%) | | | |
| mRS 0–3, n (%) | 30 (40.0) | 20 (62.5) | 0.037 |
| mRS 0–2, n (%) | 20 (26.7) | 16 (50.0) | 0.026 |

Values are presented as mean ± SD or n (%). P-values were calculated using Welch's t-test for continuous variables, Fisher's exact test for binary categorical variables and the $\chi^2$ test for categorical variables with more than two categories. ACA: Anterior cerebral artery, ICA: Internal carotid artery, ICH: Intracerebral haemorrhage, MCA: Middle cerebral artery, mRS: Modified Rankin Scale, SD: Standard deviation, WFNS: World Federation of Neurosurgical Societies.

**Table S2. Odds ratios of the admission-only prognostic models**

| Predictor | Coding/unit | mRS 0–3 OR (95% CI) | P-value | mRS 0–2 OR (95% CI) | P-value |
|---|---|---|---|---|---|
| Age | Per 10-year increase | 0.647 (0.460–0.909) | 0.0092 | 0.551 (0.385–0.758) | 0.0001 |
| Pre-morbid mRS | ≥ 2 vs. 0–1 | 0.127 (0.015–0.738) | 0.0202 | 0.054 (0.0004–0.535) | 0.0037 |
| WFNS grade | IV–V vs. I–III | 0.045 (0.012–0.125) | < 0.0001 | 0.050 (0.016–0.140) | < 0.0001 |
| Modified Fisher grade | 3–4 vs. 1–2 | 0.333 (0.114–0.934) | 0.0367 | 0.445 (0.167–1.172) | 0.1042 |
| ICH | Present vs. absent | 0.884 (0.295–2.761) | 0.8280 | 0.936 (0.298–3.147) | 1.0000 |

ORs are shown for favourable outcomes. The mRS 0–2 model was fitted using Firth's penalised logistic regression. ICH: Intracerebral haemorrhage, mRS: Modified Rankin Scale, OR: Odds ratio, WFNS: World Federation of Neurosurgical Societies.

#### Table S3. Detailed performance of the admission-only model

| Population Outcome | Cohort | AUC | Brier score | Calibration intercept | Calibration slope |
|---|---|---|---|---|---|
| Overall mRS 0–3 | Development | 0.917 (0.870–0.959) | 0.109 (0.078–0.144) | 0.000 (−0.529–0.519) | 1.000 (0.794–1.357) |
| | Validation | 0.868 (0.773–0.940) | 0.134 (0.088–0.187) | 0.587 (−0.059–1.360) | 0.827 (0.516–1.284) |
| Overall mRS 0–2 | Development | 0.920 (0.876–0.962) | 0.107 (0.077–0.138) | −0.044 (−0.575–0.471) | 1.076 (0.831–1.508) |
| | Validation | 0.870 (0.778–0.941) | 0.147 (0.097–0.205) | 0.546 (−0.077–1.214) | 0.787 (0.506–1.175) |
| Age ≥ 70 years mRS 0–3 | Development | 0.914 (0.839–0.963) | 0.122 (0.074–0.174) | −0.145 (−0.893–0.619) | 1.036 (0.748–1.608) |
| | Validation | 0.842 (0.667–0.956) | 0.169 (0.087–0.275) | 0.446 (−0.644–1.701) | 0.673 (0.271–1.245) |
| Age ≥ 70 years mRS 0–2 | Development | 0.878 (0.773–0.952) | 0.112 (0.072–0.164) | −0.286 (−1.089–0.535) | 1.053 (0.593–1.816) |
| | Validation | 0.844 (0.658–0.953) | 0.192 (0.107–0.295) | 0.381 (−0.579–1.423) | 0.577 (0.214–1.236) |

The performance metrics were calculated using the probabilities predicted from the admission-only model. The outcomes were assessed at 90 days. The development cohort comprised patients treated during 2021–2023, and the temporal validation cohort comprised patients treated during 2024–2025. The sample sizes were 160 and 85 in the overall development and temporal validation cohorts, respectively, and 75 and 32 in the corresponding cohorts among patients aged ≥ 70 years. In patients aged ≥ 70 years, coefficients derived from the overall development cohort were applied without refitting. Values in parentheses indicate bootstrap bias-corrected 95% confidence intervals based on 2000 bootstrap resamples for AUC, Brier score, calibration intercept, and calibration slope. AUC: Area under the receiver operating characteristic curve, BC: Bias-corrected, CI: Confidence interval, mRS: Modified Rankin Scale.

**Table S4. Exploratory external assessment in an independent published cohort**

| Cohort | Outcome | Favourable events | AUC | Brier score | Calibration intercept | Calibration slope |
|---|---|---|---|---|---|---|
| Independent published cohort (n = 198) | mRS 0–3 | 115 | 0.790 | 0.194 | −0.365 | 0.656 |
| | mRS 0–2 | 105 | 0.775 | 0.202 | −0.379 | 0.633 |

The premorbid mRS score was unavailable in the independent published cohort and was therefore assumed to be < 2 for all patients. The 6-month mRS was used because the 90-day mRS was unavailable. Accordingly, this analysis should be interpreted as an exploratory external assessment rather than a formal external validation. mRS: Modified Rankin Scale.

### Supplemental Figures

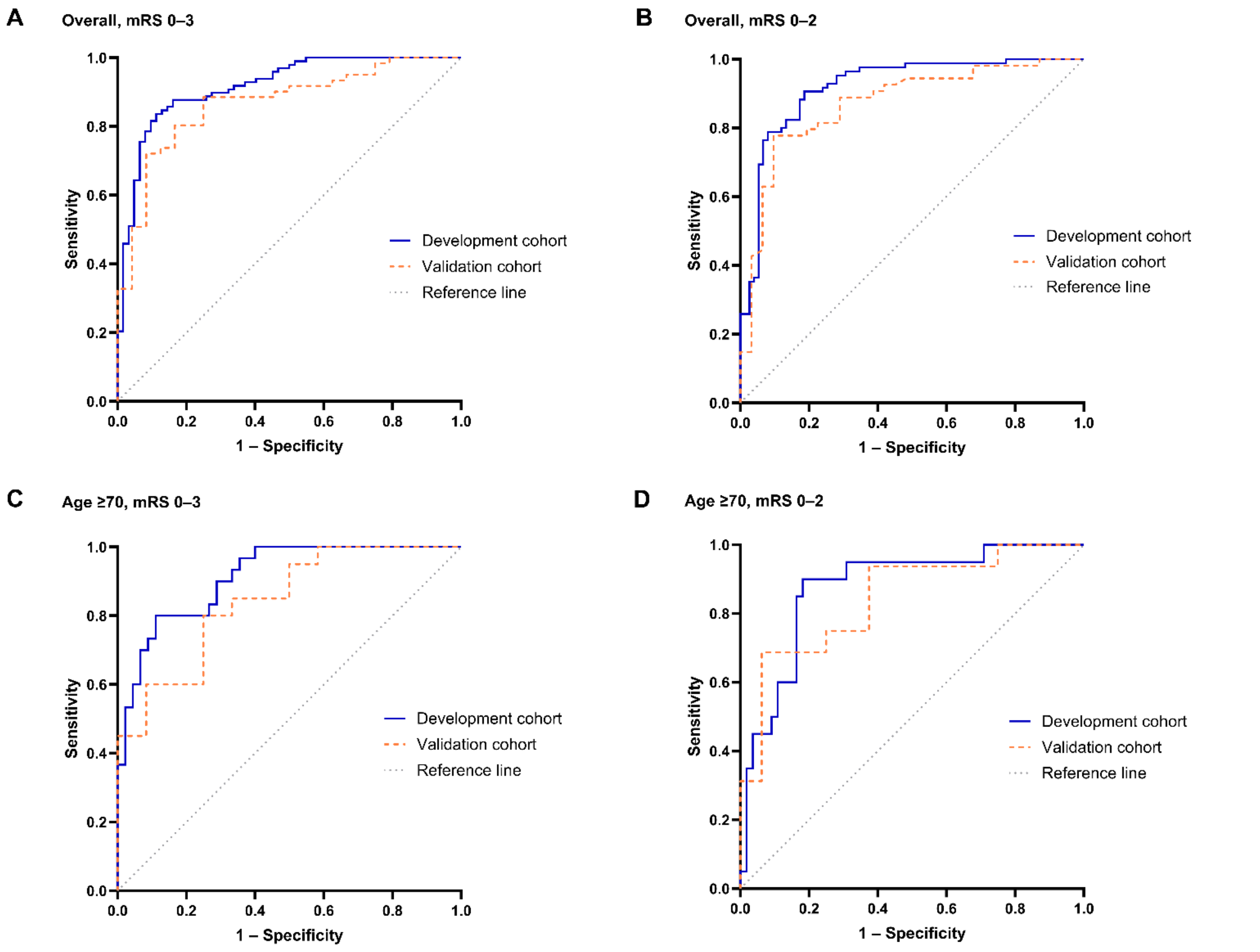


**Figure S1. Receiver operating characteristic curves of the admission-only prognostic models in the development and temporal validation cohorts.**

Receiver operating characteristic curves are shown for the overall cohort and for patients aged ≥ 70 years. (A) Overall cohort: 90-day modified Rankin Scale (mRS) score 0–3. (B) Overall cohort: 90-day mRS scores of 0–2. (C) Patients aged ≥ 70 years, with a 90-day mRS score of 0–3. (D) Patients aged ≥ 70 years with a 90-day mRS score of 0–2. The AUC values and bootstrap bias-corrected 95% confidence intervals are summarised in Table S3. AUC: Area under the receiver operating characteristic curve, CI: Confidence interval, mRS: Modified Rankin Scale, ROC: Receiver operating characteristic.

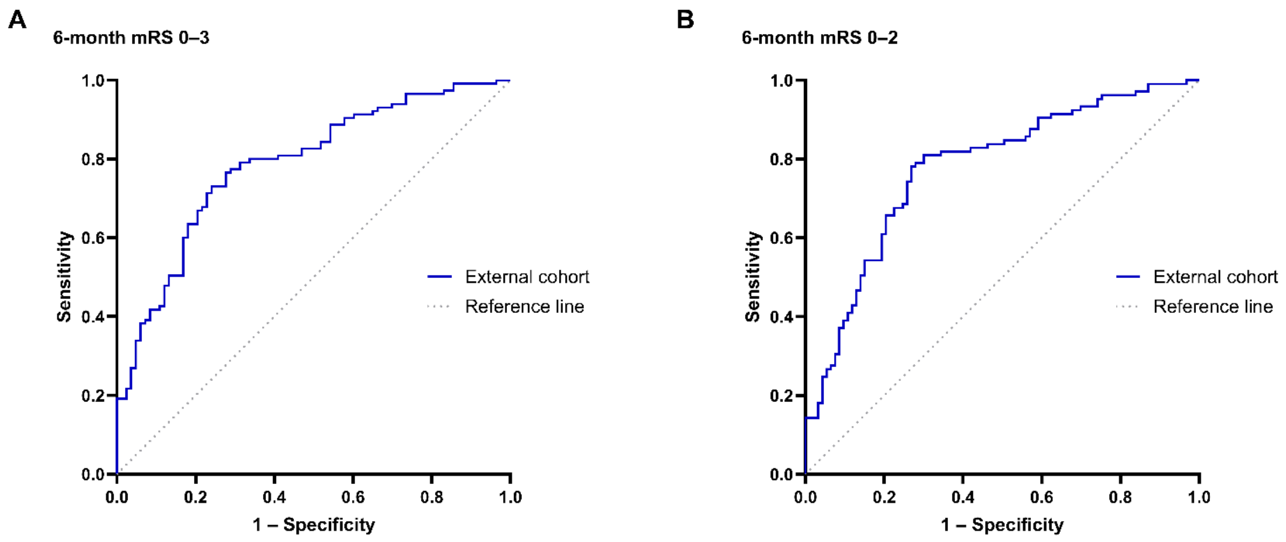


**Figure S2. Receiver operating characteristic curves for exploratory external assessment in an independent published cohort.**

ROC curves for favourable 6-month outcomes defined as mRS 0–3 (A) and mRS 0–2 (B), with AUCs of 0.790 and 0.775, respectively. The premorbid mRS score was unavailable and was assumed to be < 2 for all patients. The 6-month mRS was used because the 90-day mRS was unavailable. AUC: Area under the receiver operating characteristic curve, mRS: Modified Rankin Scale, ROC: Receiver operating characteristic.
